# Dosing interval strategies for two-dose COVID-19 vaccination in 13 low- and middle-income countries of Europe: health impact modelling and benefit-risk analysis

**DOI:** 10.1101/2021.11.27.21266930

**Authors:** Yang Liu, Carl AB Pearson, Frank G Sandmann, Rosanna C Barnard, Jong-Hoon Kim, CMMID COVID-19 Working Group, Stefan Flasche, Mark Jit, Kaja Abbas

## Abstract

**Background:** In settings where the COVID-19 vaccine supply is constrained, extending the intervals between the first and second doses of the COVID-19 vaccine could let more people receive their first doses earlier. Our aim is to estimate the health impact of COVID-19 vaccination alongside benefit-risk assessment of different dosing intervals for low- and middle-income countries of Europe.

**Methods:** We fitted a dynamic transmission model to country-level daily reported COVID-19 mortality in 13 low- and middle-income countries in the World Health Organization European Region (Albania, Armenia, Azerbaijan, Belarus, Bosnia and Herzegovina, Bulgaria, Georgia, Republic of Moldova, Russian Federation, Serbia, North Macedonia, Turkey, and Ukraine). A vaccine product with characteristics similar to the Oxford/AstraZeneca COVID-19 (AZD1222) vaccine was used in the base case scenario and was complemented by sensitivity analyses around efficacies related to other COVID-19 vaccines. Both fixed dosing intervals at 4, 8, 12, 16, and 20 weeks and dose-specific intervals that prioritise specific doses for certain age groups were tested. Optimal intervals minimise COVID-19 mortality between March 2021 and December 2022. We incorporated the emergence of variants of concern into the model, and also conducted a benefit-risk assessment to quantify the trade-off between health benefits versus adverse events following immunisation.

**Findings:** In 12 of the 13 countries, optimal strategies are those that prioritise the first doses among older adults (60+ years) or adults (20-59 years). These strategies lead to dosing intervals longer than six months. In comparison, a four-week fixed dosing interval may incur 10.2% [range: 4.0% - 22.5%; n = 13 (countries)] more deaths. There is generally a negative association between dosing interval and COVID-19 mortality within the range we investigated. Assuming a shorter first dose waning duration of 120 days, as opposed to 360 days in the base case, led to shorter optimal dosing intervals of 8-12 weeks. Benefit-risk ratios were the highest for fixed dosing intervals of 8-12 weeks.

**Interpretation:** We infer that longer dosing intervals of over six months, which are substantially longer than the current label recommendation for most vaccine products, could reduce COVID-19 mortality in low- and middle-income countries of WHO/Europe. Certain vaccine features, such as fast waning of first doses, significantly shorten the optimal dosing intervals.

**Funding:** World Health Organization

## Introduction

By October 2021, 22 COVID-19 vaccines were in use globally.^1^ However, vaccine supply has struggled to meet global demand. In particular, low- and middle-income countries (LMICs) faced delays in vaccine roll-out. These constraints are expected to ease in late 2022 as additional production capacity becomes available.^2^ In the interim, it is vital that countries can maximise the health impact of the available vaccine supplies based on context-specific COVID-19 epidemiology and SARS-CoV-2 transmission dynamics, pre-existing immunity, and COVID-19 vaccine safety, immunogenicity, and efficacy.^3^

Most of the vaccines currently available involve two doses with recommended between-dose intervals as tested in the clinical trials. These dosing intervals are generally 3-4 weeks, although they may differ by vaccine product. However, in practice, countries may use dosing intervals longer than recommended due to a wide range of factors, including but not limited to administrative and logistic constraints (e.g. vaccine clinic capacity), vaccine shortages, and the comparable and potentially higher vaccine efficacy using certain extended dosing intervals.^4^ Particularly, countries need to consider the trade-off between partial protection (induced by one dose) of a larger number of individuals versus full protection (induced by two doses) of fewer individuals. However, there is limited evidence on the population-level health impacts related to these trade-offs.^5^ Moreover, there are concerns about the benefits of the COVID-19 vaccine versus the harms from adverse events following immunisation (AEFI) depending on the age of the vaccinated individuals, which may change conclusions of the overall health benefits of vaccination.

To address this evidence gap, we used a mathematical modelling approach that incorporated two-dose dynamics to assess the health impacts (i.e. COVID-19 mortality) of different dosing intervals in the LMICs in the World Health Organisation (WHO) European Region between March 2021 and December 2022. We aim at identifying the optimal dosing interval strategy. The evidence generated could inform COVID-19 vaccine policies as vaccines roll out - rapid shifts in policies have been shown feasible.^6^ As the LMICs within the Region represent a wide range of population age structures and epidemic histories; the evidence could be valuable to vaccine policies in LMICs elsewhere in the world facing similar issues.

We explored dosing interval policy approaches: (1) by setting fixed dosing intervals of different lengths; (2) by setting coverage targets (e.g. to first cover the entire adult population with the first dose). In the baseline scenario, we based the vaccine characteristics on those of the Oxford/AstraZeneca COVID-19 vaccine AZD1222 (hereafter AZD1222) and followed up with sensitivity analyses for vaccine characteristics similar to other COVID-19 vaccines.

## Methods

### The mathematical modelling process

We adapted CovidM, an existing transmission dynamic model of SARS-CoV-2, which has already been extensively described elsewhere,^7–12^ to estimate the health impact of COVID-19 vaccination. We attach the complete parameter table (relevant for the entire Methods), together with their references, as Supplemental Table S1.

In brief, it is an SEIR-type (susceptibles, exposed, infectious, and recovered) compartmental model consisting of 16 age groups (0-4 to 75+ years with five-year increments) to incorporate age-specific susceptibility, clinical fractions, population sizes, and contact patterns. Within each age group, 13 compartments (Figure 1) were used to capture the population dynamics.

**Figure 1.**
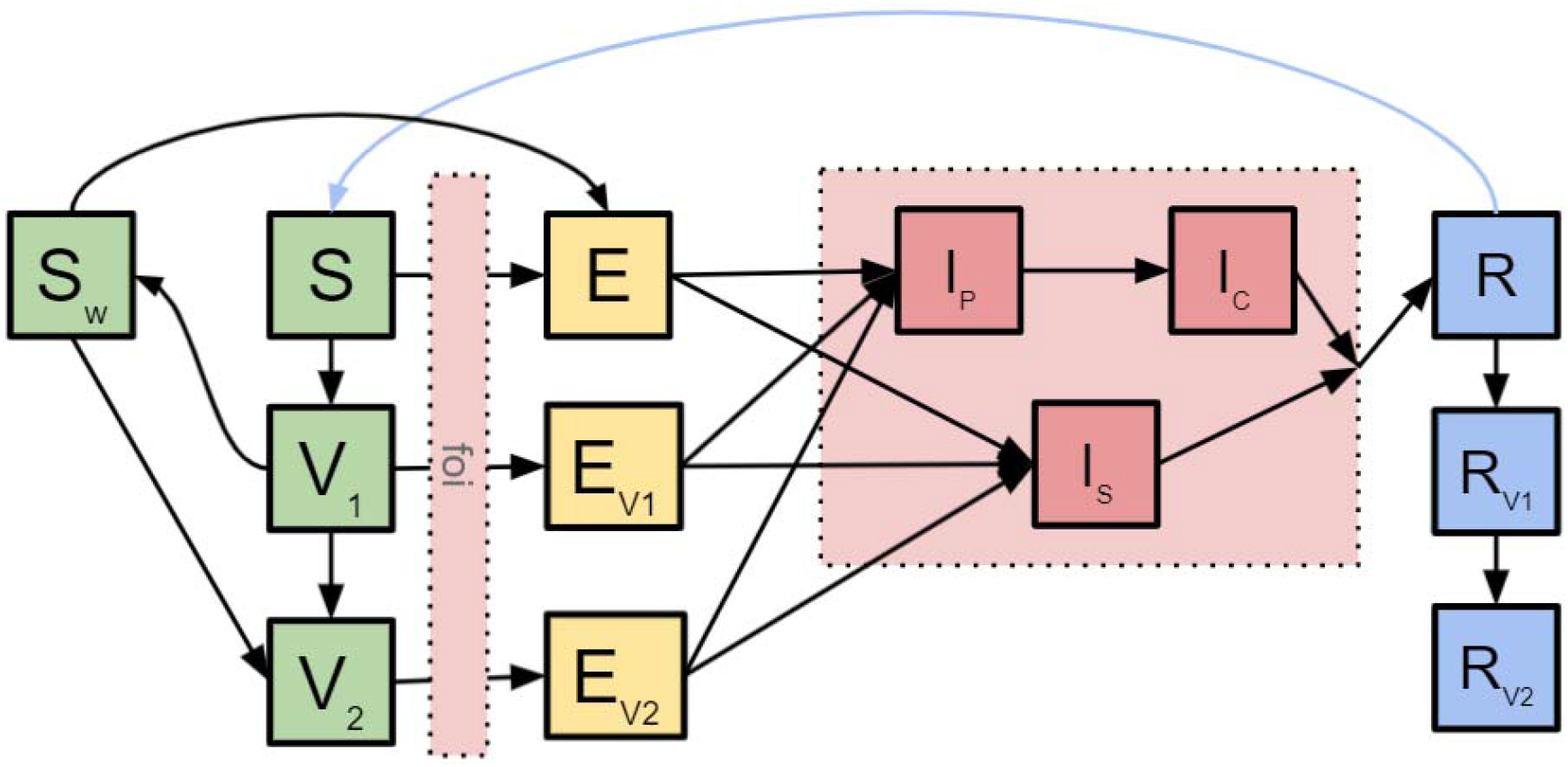
SARS-CoV-2 transmission dynamics and COVID-19 vaccination impact. The conceptual diagram describes the underlying mathematical models of SARS-CoV-2 transmission dynamics and COVID-19 vaccination impact. S - susceptible; V_1_ - individuals protected by the first dose only; V_2_ - individuals protected by both doses; S_w_ - individuals who have received their first dose but the protection has waned; E - exposed; E_v1_ - exposed progressed from individuals in V_1_; E_v2_ - exposed progressed from individuals in V_2_; I_p_ - pre-clinical infectious individuals; I_c_ - clinical individuals; I_s_ - subclinical individuals; R - recovered; R_v1_ - previously infected individuals whose infection-induced immunity has yet to wane and who have received the first dose; R_v2_ individuals whose infection-induced immunity has yet to wane and who have received both doses.

Compared to a classic SEIR model, this framework has several unique features. First, individuals protected by one and two doses are modelled separately (V_1_ and V_2_ in Figure 1), allowing for the incorporation of dose-specific dynamics and parameters. Second, we accounted for potential waning among those who have only received their first doses (S_w_ in Figure 1), as suggested by evidence.^4,13^ Third, recovered individuals could receive vaccinations (R_v1_ and R_v2_ in Figure 1). In this region, antibody tests have not been used to qualify individuals for vaccination. The model framework has left out breakthrough infections among those with a COVID-19 infection history who had been vaccinated, capturing the potential boosting effects vaccines may have among recovered individuals.^14^ Fourth, to account for the significant deviation from the public’s normal routine, we used Google Community Mobility Index (hereafter “mobility index”) to adjust the contact matrices.^15^ This scaling method has been previously presented elsewhere.^16^ A brief description can be found in Supplemental Table S2 and Supplemental Methods p16.

This model was fitted to the country-level daily reported COVID-19 deaths before March 2021 in 13 LMICs in the WHO European Region: Albania, Armenia, Azerbaijan, Belarus, Bosnia and Herzegovina, Bulgaria, Georgia, Republic of Moldova, Russian Federation, Serbia, North Macedonia, Turkey, and Ukraine for three variables: infection introduction date, basic reproduction number, and under-reporting rate. Country-specific fitted results can be found in the GitHub repository attached to this study. We did not fit the model to the remaining seven countries due to data availability (n = 1), data sparsity issues (<10 deaths/ day throughout the fitting period, n = 4) or significant changes in ways tallying COVID-19 mortality (n = 2). Country characteristics are captured by population age structures, age-specific contact patterns, changes in the mobility index, epidemic history, and immunity levels prior to vaccine introduction. The fitting approach was based on maximum likelihood estimation and differential evolution optimisation and has been described in detail elsewhere.^16^

Given the relatively short time horizon of this study, we modelled COVID-19 severe cases and mortality as processes, apart from the population dynamics presented in Figure 1 (i.e., there is no removal of individuals from the modelled population). The total COVID-19 severe case incidence and mortality is the product of age-specific infection counts, infection-hospitalisation risks/infection-fatality risks, and functions describing the delay between infection and hospitalisation/ mortality. Equations describing these processes can also be found in the Supplemental Methods p15.

### Vaccine characteristics

In the base case scenario, we considered a vaccine product with characteristics similar to those of AZD1222 and incorporated five types of vaccine effects (i.e. Infection-, disease-, severe case-, mortality-, and onward transmission-reducing) into the model (Figure 2, a). For the rationale behind specific parameters used, please refer to Supplemental Table S3.^11^

**Figure 2.**
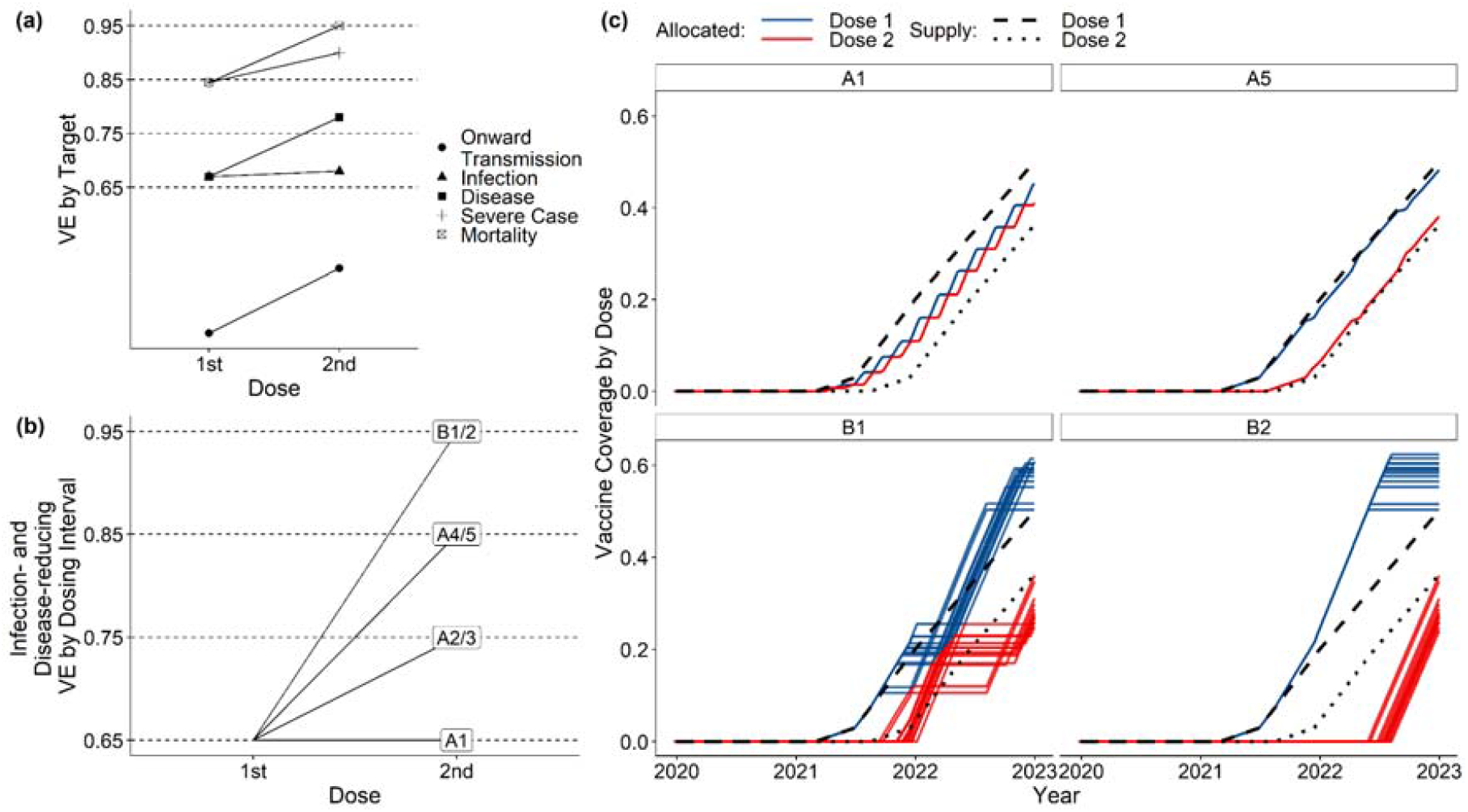
COVID-19 vaccine and vaccination program-related parameters. (a) Vaccine efficacy by target, respectively describing onward transmission blocking, infection-, disease-, severe case- and mortality-reducing vaccine efficacies. (b) Sensitivity analysis parameter set that describes a condition where longer dosing interval is associated with larger incremental change incurred by the second doses in infection- and disease-reducing vaccine efficacies. (c) Vaccine supply and allocation conditions by dose. We assume a 24-week delay between the supply of first and second doses. We did not show strategies A2, A3 and A4 as they reflect incremental changes between A1 and A5. Each line represents a country. Refer to Table 1 for descriptions of vaccination strategies.

Given the substantial uncertainty around the estimates of vaccine efficacies and the possibility that a country may use other (or multiple) vaccine products, we conducted sensitivity analyses over four dimensions around the values for infection- and disease-reducing vaccine efficacies for the first and second doses (Supplemental Methods p17). There is also limited evidence that suggests higher vaccine efficacy after the second dose when the dosing interval is longer than label recommendations.^5^ We explored how this observation may affect the optimal dosing strategy (Figure 2, b).

**Table 1.** Vaccination strategies. Definitions of vaccine dosing strategies. Older adults: those above 60 years of age; young adults: those between 20 and 59 years of age.

| Strategy | Description |
| --- | --- |
| A1 | Individuals will receive their 2nd doses **4** weeks after their first doses. |
| A2 | Individuals will receive their 2nd doses **8** weeks after their first doses. |
| A3 | Individuals will receive their 2nd doses **12** weeks after their first doses. |
| A4 | Individuals will receive their 2nd doses **16** weeks after their first doses. |
| A5 | Individuals will receive their 2nd doses **20** weeks after their first doses. |
| B1 | Older adults will receive their 2nd doses after all older adults receive their first doses. Young adults will receive their 1st doses after older adults uptake targets are reached. Young adults will only receive 2nd doses when all young adults have received their 1st doses. |
| B2 | Older adults will receive their 2nd doses after all individuals (older adults + young adults) receive their first doses; young adults will receive their 2nd doses after older adults uptake targets are reached, and all young adults have received their first dose |

We implemented a 14-day delay between vaccination (i.e. doses administered) and immunisations (i.e. protection developed).

There is evidence showing the potential waning of the first dose when the second dose has not been administered in a timely manner.^4,13^ In this study, we assume this waning process (loss of protection from the vaccine) occurs on average over 360 days. In case this waning process occurs faster than expected, a shorter waning duration of 120 days was tested as a worst-case scenario sensitivity analysis.

### Strategies regarding dosing intervals

In this study, we used two general approaches to set dosing intervals (Table 1). Some countries may choose to fix an *n* week dosing interval between the first and second doses for all individuals. Other countries may aim to cover all individuals with the first dose before vaccinating anyone with the second dose. We expanded these two general approaches into seven dosing interval strategies (Figure 2, c). We assume protection following the first dose may wane before the second dose is given; protection following the second dose, however, is assumed not to wane regardless of the dosing interval.

### Other Assumptions around the Vaccination Programmes

We have previously shown that an age-based vaccine prioritisation strategy that targets older adults first (i.e. 60+) and then moves onto young adults (i.e. 20-59) consistently performs comparably or better than other age-based vaccine prioritisation strategies.^16^ All dosing intervals we tested were in addition to this age-based vaccine prioritisation strategy. We additionally assume the uptake goal for older adults is 90% and for younger adults 70%. In other words, when the uptake level has reached 90% among those above 60 years, the vaccination program is considered complete for that age group and will move on to the next age group. These two uptake thresholds are feasible based on age-specific uptake levels in countries with relatively fast vaccine roll-out in older adults.

We assumed the starting date of vaccination programs to be 01 March 2021 in LMICs from WHO press releases. Based on the COVAX vaccine supply forecast to inform vaccine supply,^2^ we set the vaccine supply targets of the first doses to be 3% of the total population by mid-2021 and 20% of the total population by the end of 2021. With a slightly faster rate, we assume this program will aim at covering 50% of the population with their first dose by the end of 2022. This roll-out rate is in line with the slowest adaptors among LMICs within the Region (Supplemental Figure S1). To investigate the impacts of vaccine supply delay, we assume the second dose will become available 24 weeks after their initial inoculation dates.

As vaccines roll out, contact levels within the population may recover. In this study, we assume contacts gradually recover to near pre-pandemic levels (i.e. 90% recovery) over a year’s time from March 2021 following a sigmoid function. We do not account for reactive non-pharmaceutical interventions in response to surging infections as the action threshold (i.e. the definition of “surging infections”), and the action intensity (e.g. a lockdown or a face mask mandate) may vary significantly by country.

### Consideration around variants of concern

The emergence of variants of concern (VOCs) may reduce the efficacy of vaccines. We investigated the potential effects of VOC emergence by modifying disease dynamics and vaccine efficacy. To capture the potential increase in transmissibility, we implemented a 50% increase in transmissibility. To capture the potential increase in severity, we implemented 50% increases in infection fatality and hospitalisation regardless of vaccination status. To account for potential immune escape, we included a 40% reduction in infection-blocking vaccine efficacy. These characteristics have been assumed based on current evidence around the Delta variant.^11,17^ All three changes described were introduced into the simulation processes on 15 April 2021 simultaneously, broadly aligning with the approximate timing of the large-scale emergence of the Delta variant in western Europe.^18,19^

### Criteria for optimal dosing strategy

We used the cumulative mortality between 01 March 2021 and 31 December 2022 COVID-19 as the primary decision-making metric. The optimal dosing interval strategy minimises the cumulative mortality. As discussed above, COVID-19 mortality is the product of age-specific infection counts, infection-fatality ratios, and a temporal delay function representing the interval between becoming infected and dying of COVID-19 (Supplemental Methods p15). We compare dosing interval strategies by calculating the percentage difference in cumulative mortality relative to the reference strategy B1.

### Benefit-risk analysis of COVID-19 vaccination versus adverse events following immunisation

In line with the previous parameterisation based on AZD1222, we quantified the risk of fatal AEFI based on major thromboembolic events (blood clots) with low platelet count (thrombocytopenia) as reported for the AZD1222. In the UK, up to 01 September 2021, there have been 416 cases in total, of which 45 cases occurred after the second dose.^20^ A total of 72 deaths occurred, with 6 deaths reported after the second dose. Given the different number of doses given by age in the UK, the age-specific gradient of the risk of developing these serious AEFIs is about 20.5 per 1 million doses in individuals aged 18-49 years and 10.9 per 1 million doses in individuals aged ≥ 50 years after the first dose; and 0.9 per 1 million doses and 1.9 per 1 million doses after the second dose, respectively.^20^

Based on the results of the different dosing intervals in the main analysis, we used these age-specific rates of occurrence after the first and second dose, respectively, times the proportion of fatalities after the first and second dose to estimate the total number of fatal AEFIs based on the number of vaccine doses used in each country.

We then traded off the age-specific mortality from COVID-19 versus the age-specific mortality caused by AEFIs. We took the worst-performing dosing strategy in terms of mortality as the baseline to estimate the benefit-risk ratios, which quantify the benefit of additional deaths prevented by the vaccines versus the harm from additional deaths caused by AEFIs. For this analysis, strategy A1 is taken as the baseline to obtain positive results for the benefits (and the benefit-risk ratios) to facilitate the interpretation of results. If one was to consider a no-vaccination strategy (which is highly unlikely for COVID-19 in general, and in particular for the countries included here), then all of the benefit-risk ratios against no-vaccination are expected to be greater than 1.0 given the estimated much larger rate of outcomes in SARS-CoV-2 positive cases among unvaccinated individuals than vaccinated individuals.^21^

All the program code and data used in the study are publicly accessible online at https://github.com/yangclaraliu/COVID_Vac_Delay.

### Role of the funding source

The funders were involved in study design, data collection, data analysis, data interpretation, writing of the report, and the decision to submit for publication. We had discussions and received feedback from the members of the WHO Strategic Advisory Group of Experts on Immunization (SAGE) Working group on COVID-19 vaccine impact modelling and Immunisation, and Vaccines related Implementation Research Advisory Committee (IVIR-AC). All authors had full access to all the data in the study and had final responsibility for the decision to submit for publication. This study was approved by the ethics committee (Ref 26318) of the London School of Hygiene & Tropical Medicine.

## Results

### The implication of different dosing interval strategies

With a 24-week delay in vaccine supply, under strategies A1 and A5, individuals were able to receive their second doses at the dosing interval prescribed (i.e. 4-20 weeks). Under strategies B1 and B2, the second doses are not provided based on a fixed dosing interval - but on conditions that coverage targets have been met in target populations. The mean dosing intervals under strategy B1 range between 24 and 29 weeks and the median between 22 and 34 weeks. The mean dosing intervals under strategy B2 range between 45 and 57 weeks and the median between 43 and 56 weeks (Figure 3). Under strategy B1, dosing intervals are bi-modally distributed, with the first peak representing the dosing intervals of older adults and the second peak representing the dosing intervals of younger adults. The mean dosing interval among older adults under strategy B1 is comparable to that under strategy A5 in most countries and even strategy A4 in some countries (e.g. Azerbaijan and Turkey).

**Figure 3.**
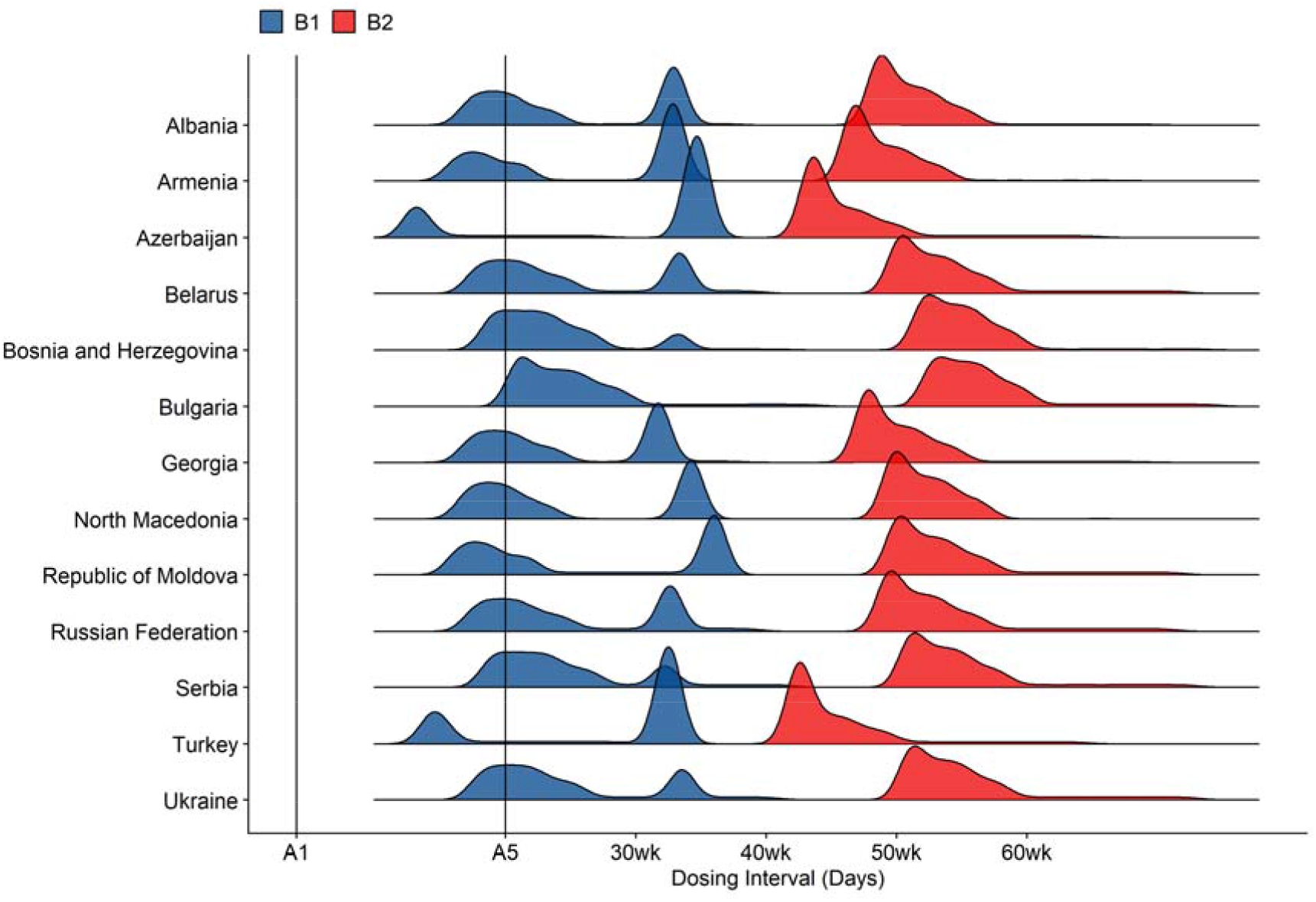
COVID-19 dosing intervals under strategies B1 and B2. These dosing strategies do not prescribe fixed dosing intervals. Vaccine allocations depend on whether or not coverage goals have been met in certain target groups. The distributions are outputs from dose allocation algorithms that capture such conditional relationships. Refer to Table 1 for descriptions of vaccination strategies.

### Optimal strategies under the base case scenario

At the beginning of the simulated vaccine roll-out processes (i.e. 01 March 2021), from model fitting, we estimated immunity levels in 13 LMICs in the WHO European region to range from 1.22% to 64.6%.

Assuming VOC emergence and a mean first-dose waning duration of 360 days, B1 and B2 were optimal strategies for minimising COVID-19 mortality in 12 of 13 countries investigated and were comparable with each other, with the exception of Serbia (Figure 4, a). The model fitting process predicted that Serbia has the smallest proportions of the population who remained susceptible by 01 March 2021.Strategy A1 may be associated with on average 10.2% higher cumulative mortality between 01 March 2021 and 31 December 2022 [range: 4.0% - 22.52%; n = 13 (countries)]. There is a negative association between dosing interval and relative cumulative mortality.

**Figure 4.**
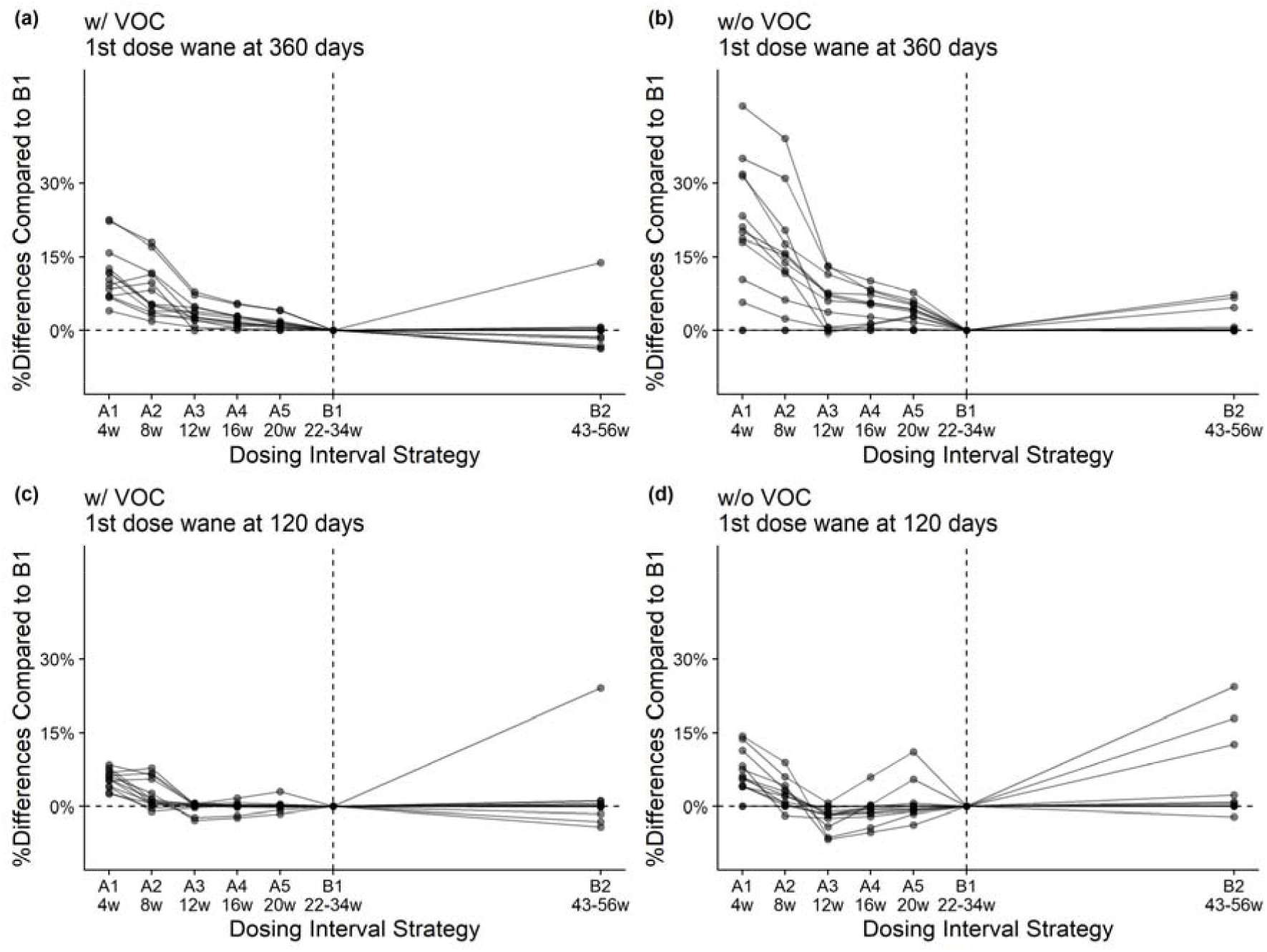
Percentage difference for different vaccination strategies relative to strategy B1. Each line represents a country. Detailed descriptions of the vaccine dosing strategies can be found in Table 1. Mean dosing intervals are presented as the second rows of x-axes labels (unit = weeks). Results from sensitivity analyses around the first dose waning durations (360 days vs 120 days) and the emergence of variants of concern (VOCs) are also presented.

Without VOC emergence, the advantages of strategy B1 are more evident, although B1 and B2 are still broadly comparably (Figure 4, b). The negative association between dosing interval and COVID-19 mortality is also observed. However, strategy B2 is worse off than B1 in Albania, Azerbaijan, and Turkey by more than 5% (using the mortality under B1 as the denominator). Strategy A1 is associated with an average of 15.9% increase in cumulative mortality between 01 March 2021 and 31 December 2022 [range: 0% - 45.7%; n = 13]. Note that in countries where no additional outbreak occurs during this period, the proportional difference between any strategies relative to strategy B1 is 0.

Assuming a mean first-dose waning duration of 120 days, the superiority of strategy B1 is no longer evident. Optimal dosing strategy shifts towards the range of A2 to A4 (8 to 16 weeks, Figure 4, c-d).

### Sensitivity analyses around vaccine efficacy

There is still considerable uncertainty around the relationship between dosing interval and the vaccine efficacy achievable after the second dose of the COVID-19 vaccines.^22^ In this study, we examined the extreme case where vaccine efficacy incrementally increases as dosing interval gets longer (Figure 1, b). We fixed the first dose infection-reducing vaccine efficacy at 65% to make it comparable to the results from the base case scenarios.

With VOC emergence, the comparative advantage of strategy B1 and B2 compared to strategies A1-A5 becomes slightly higher (Supplemental Figure S2). Without VOC emergence, however, the percentage differences in cumulative COVID-19 mortality compared to strategy B1 remains broadly consistent.

While varying four dimensions of vaccine efficacy (i.e. first and second doses infection- and disease-reducing effects), we found that strategies B1 and B2 were comparable (within 5% difference) and advantageous for all countries except for Serbia, consistent with the baseline results (Supplemental Figure S3).

### Benefit-risk analysis

The additional deaths prevented by dosing strategies compared to the reference strategy A1 was always greater than the additional deaths due to fatal adverse events caused by these strategies. Most benefit-risk ratios of vaccination strategies in comparison to reference strategy A1 were greater than the additional harms (97.8%; n=178/182, with and without VOC combined); in four ratios, the benefits of strategy A2 or B2 were smaller than those of strategy A1 (A2 in Bosnia and Herzegovina, Serbia and Ukraine and B2 in Serbia). The additional benefits for strategies A2 and A3 were much greater than the additional harms among all other vaccination strategies, favouring a dosing interval of 8-12 weeks (compared to 4 weeks). The dominant benefit of strategy A3 was even more pronounced in the absence of VOC emergence (Figure 5, B).

**Figure 5.**
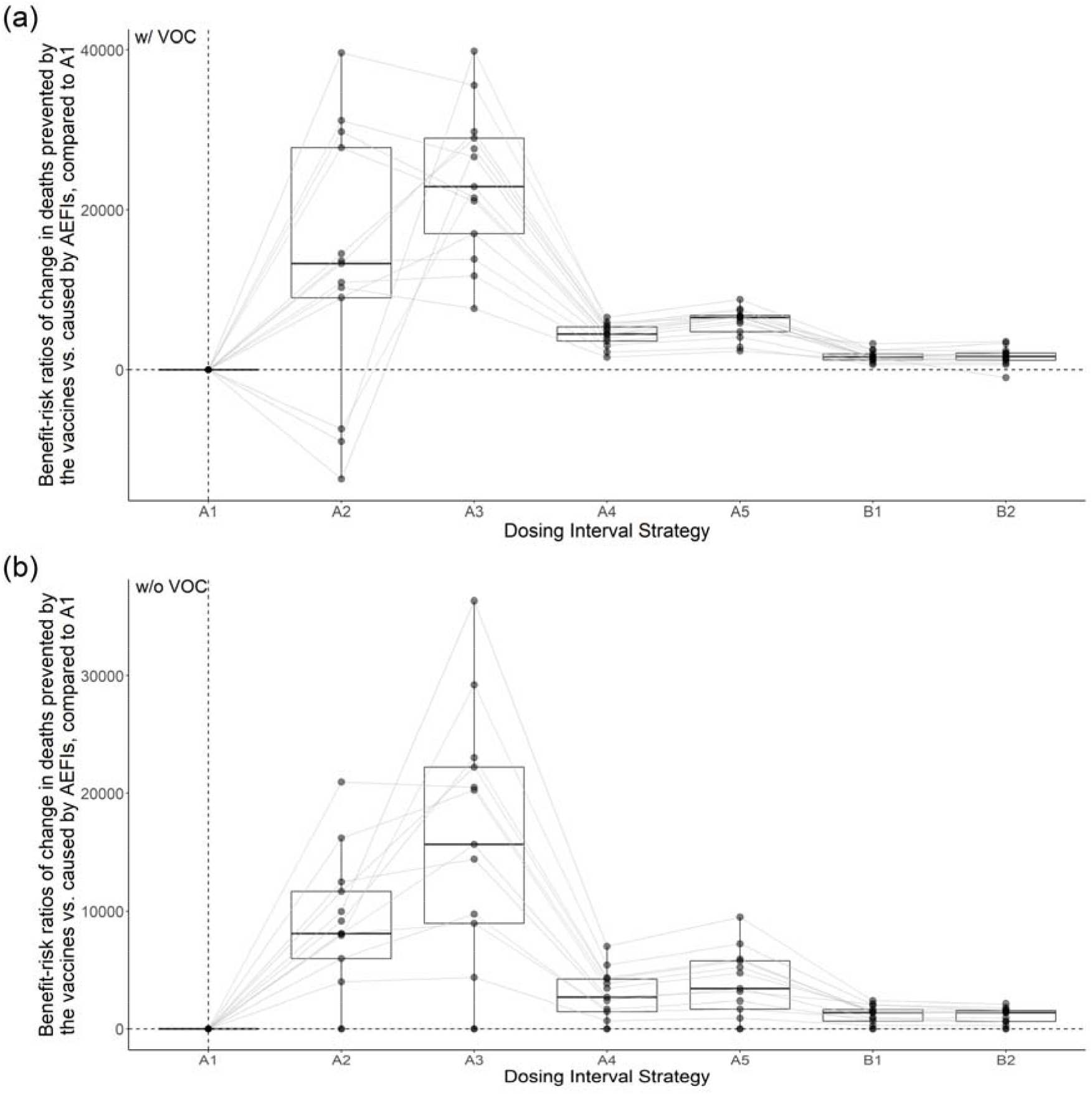
Benefit-risk ratios of vaccination strategies. Benefit-risk ratios of the different dosing interval strategies in comparison to strategy A1 are presented. Strategies A1-A5 and B1-B2 are arranged broadly based on mean effective dosing intervals. Each line represents one country. The benefit-risk ratios quantify the change in deaths prevented by the vaccines versus caused by AEFIs as compared to strategy A1. Refer to Table 1 for descriptions of vaccination strategies.

Although we did not include the health and societal benefits of avoiding non-fatal SARS-CoV-2 positive cases, the general trend of our estimated outcomes (Supplemental Figure 3) follows a similar trend as mortality, which indicates that including additional benefits should result in similar findings as shown in Figure 5.

## Discussion

We explored the health implications of different COVID-19 vaccine dosing interval scenarios in 13 low- and middle-income countries of the WHO European Region. We explored seven strategies that involved either fixed dosing intervals of between 4 weeks and 20 weeks or a conditional policy that depended on vaccination goals being met. We found that vaccinating all older adults or adults with the first dose (strategy B1 and B2) resulted in the lowest COVID-19 cumulative mortality for 12 of the 13 countries, while a dosing interval of 4 weeks (strategy A1) resulted in the largest number of cumulative mortality. Small cumulative mortality was associated with an early increase in the proportion of the population vaccinated with the first dose. The results we have observed are robust to different vaccine characteristics assumptions of vaccine efficacy estimates versus dosing interval relationships.

There is a negative association between dosing intervals and cumulative mortality when mean dosing intervals range between 4 and 26 weeks. This association may not persist with even longer dosing intervals of 43-56 weeks (as in strategy B2). While longer-than-currently recommended dosing intervals may be beneficial in reducing COVID-19 mortality, exceptionally long dosing intervals should be cautioned against. A faster waning of the protection derived by the first dose (with a mean of 120 days, instead of 360 days in the base case scenario) made strategies with shorter, fixed dosing intervals (i.e. A2-4) more favourable.

When trading off the benefits of the vaccines in preventing deaths versus the harm from deaths caused by adverse events following immunisation, we found that the additional benefits of the strategies compared to strategy A1 exceeded the additional risks for the vast majority of dosing interval scenarios. The ratio of benefits to risks was highest for strategies A2-A3 (intervals of 8-12 weeks), following the age-dependent rates of (prevented) deaths from COVID-19 and adverse events. Including additional outcomes and benefits for society is expected to lead to similar results.

Related studies in high-income country settings of Canada, Netherlands, the United Kingdom, and the United States also found that delaying the second dose of a two-dose COVID-19 vaccine series is beneficial.^23–26^ Moghadas et al. used an agent-based model to compare two strategies of either vaccinating more individuals with the first dose and delaying the second dose or administering the 2-dose series according to the recommended dose spacing for Pfizer-BioNTech (BNT162b2) and Moderna (mRNA-1273) vaccines.^23^ They suggested that depending on pre-existing immunity levels, additional hospitalisations and deaths could be averted by delaying the second dose due to vaccine prioritisation of individuals at higher risk of severe outcomes. Romero-Brufau et al. conducted a similar study to assess the cumulative public health impact over 6 months for delaying the second dose of Pfizer-BioNTech and Moderna vaccines and inferred that the delayed second-dose strategy for people under 65 years was a favourable strategy.^27^ Based on evidence synthesis of related studies^28–30^ and our study, we infer that for two-dose vaccines, the optimal dosing interval depends on multiple factors, including vaccine efficacy and effectiveness, waning dynamics of vaccine-induced immunity, vaccination coverage, vaccine supply rates, pre-existing naturally acquired immunity, and country-specific vaccine prioritisation plans.

In our study, we fitted a dynamic transmission model using reported daily COVID-19 mortality in 13 low- and middle-income countries of the WHO European region. We explored a wide range of dosing interval strategies of prime interest to countries that may face constraints in vaccine supply. We performed extensive sensitivity analyses around key epidemiological parameters like the emergence of variants of concern and vaccine efficacy estimates. We extended the analysis by trading off the benefits of the vaccine against the potential harm from adverse events following immunisation with the COVID-19 vaccines.

We were not able to extend the fitting window of this study further into 2021 due to data availability issues (e.g. age-specific vaccine uptake, vaccine products in-use, strain-specific test positive rates). We have not incorporated reactive non-pharmaceutical interventions (e.g. lockdown enacted due to surging infections) as the specific implications are uncertain and country-specific. We were unable to capture breakthrough infections or vaccine waning among vaccinated individuals who have experienced previous infections. These pathways are biologically sound but would make it challenging to track vaccine allocations (e.g. making sure everyone only receives two doses). Given that these mechanisms are either considered relatively rare or are not widely characterised by empirical data, we did not include them in this study.

Our study shows that a dose-specific roll-out strategy that led to an average six-month dosing interval, which is substantially longer than the current label recommendation for most vaccine products available in the European market, may be able to minimise COVID-19 mortality in the LMICs in the WHO European Region. Countries included in this study have diverse population age structures, contact patterns, and epidemic histories – the overall conclusions are valuable to COVID-19 vaccine dosing policy-making in LMICs elsewhere in the world.

## Data Availability

This study has only used publicly available data sources, which have been clearly referenced in text.

## Contributors

YL, FGS, MJ, and KA conceptualised the study. YL, CABP, and FGS developed the model and conducted the health impact assessment and benefit-risk analysis. RB compiled the evidence on vaccine efficacy estimates. JHK and SF interpreted the comparative analysis of dosing interval strategies. YL wrote the original draft, and all authors contributed to reviewing and editing of the manuscript for important intellectual content and have approved the final version.

## Declaration of interests

We declare no competing interests.

## Acknowledgements

We are grateful for the helpful discussions and feedback from Nicholas Grassly (Imperial College London), Raymond Hutubessy (World Health Organization), Sarah Pallas (Centers for Disease Control and Prevention), and members of the WHO Strategic Advisory Group of Experts on Immunization (SAGE) Working group on COVID-19 vaccine impact modeling and Immunisation and Vaccines related Implementation Research Advisory Committee (IVIR-AC). We thank Nicholas G Davies (LSHTM) for his work on CovidM upon which this analysis has been built upon.

We thank the following agencies for their support: World Health Organization (202683881, 202604060), Bill & Melinda Gates Foundation (INV-009125, INV-003174, OPP1184344), European Commission (101003688), Medical Research Council (MC_PC_19065), National Institute of Health Research (200929), Foreign, Commonwealth and Development Office (UK)/Wellcome Trust (221303/Z/20/Z), Wellcome Trust (208812/Z/17/Z). FGS and MJ were supported by the NIHR Health Protection Research Unit (HPRU) in Modelling and Health Economics, a partnership between Public Health England (PHE), Imperial College London, and the London School of Hygiene & Tropical Medicine (LSHTM; grant code NIHR200908). MJ was supported by the NIHR HPRU in Immunisation at LSHTM in partnership with PHE (grant reference code NIHR200929).

We would like to acknowledge the support we received from the CMMID Working Group members: Christopher I Jarvis, Rachael Pung, C Julian Villabona-Arenas, Katharine Sherratt, Kerry LM Wong, Simon R Procter, Katherine E. Atkins, Paul Mee, Samuel Clifford, Hamish P Gibbs, Oliver Brady, David Hodgson, James D Munday, Nikos I Bosse, Timothy W Russell, Nicholas G. Davies, Sebastian Funk, William Waites, Emilie Finch, Joel Hellewell, Sam Abbott, Adam J Kucharski, Lloyd A C Chapman, Ciara V McCarthy, Yalda Jafari, Amy Gimma, Gwenan M Knight, Alicia Rosello, Billy J Quilty, Rachel Lowe, Graham Medley, Mihaly Koltai, Matthew Quaife, Kathleen O’Reilly, Sophie R Meakin, Stéphane Hué, Akira Endo, Damien C Tully, W John Edmunds, Kiesha Prem, Rosalind M Eggo.

They are partly funded by the Bill & Melinda Gates Foundation (INV-001754: MQ; INV-003174: KP; INV-016832: SRP; NTD Modelling Consortium OPP1184344: GFM; OPP1139859: BJQ; OPP1191821: KO’R). CADDE MR/S0195/1 & FAPESP 18/14389-0 (PM). EDCTP2 (RIA2020EF-2983-CSIGN: HPG). ERC Starting Grant (#757699: MQ). ERC (SG 757688: CJVA, KEA). This project has received funding from the European Union’s Horizon 2020 research and innovation programme - project EpiPose (101003688: AG, KLM, KP, WJE). This research was partly funded by the Global Challenges Research Fund (GCRF) project ‘RECAP’ managed through RCUK and ESRC (ES/P010873/1: CIJ). HDR UK (MR/S003975/1: RME). HPRU (This research was partly funded by the National Institute for Health Research (NIHR) using UK aid from the UK Government to support global health research. The views expressed in this publication are those of the author(s) and not necessarily those of the NIHR or the UK Department of Health and Social Care200908: NIB). MRC (MR/N013638/1: EF; MR/V027956/1: WW). Nakajima Foundation (AE). NIHR (16/136/46: BJQ; 16/137/109: BJQ; 1R01AI141534-01A1: DH; NIHR200908: AJK, LACC, RME; NIHR200929: CVM, NGD; PR-OD-1017-20002: AR, WJE). Royal Society (Dorothy Hodgkin Fellowship: RL). Singapore Ministry of Health (RP). UK DHSC/UK Aid/NIHR (PR-OD-1017-20001: HPG). UK MRC (MC_PC_19065 - Covid 19: Understanding the dynamics and drivers of the COVID-19 epidemic using real-time outbreak analytics: NGD, RME, SC, WJE; MR/P014658/1: GMK). UKRI (MR/V028456/1: YJ). Wellcome Trust (206250/Z/17/Z: AJK, TWR; 206471/Z/17/Z: OJB; 208812/Z/17/Z: SC; 210758/Z/18/Z: JDM, JH, KS, SA, SFunk, SRM; 221303/Z/20/Z: MK). No funding (DCT, SH)”

## Notes

### Competing Interest Statement

The authors have declared no competing interest.

## Bibliography

1 Vaccine Centre at LSHTM. COVID-19 vaccine tracker. 2020. https://vac-lshtm.shinyapps.io/ncov_vaccine_landscape/ (accessed May 18, 2021).

2 CEPI, Gavi, UNICEF, WHO. COVAX Global Supply Forecast. 2021; published online Sept 8. https://www.gavi.org/news/document-library/covax-global-supply-forecast (accessed Sept 16, 2021).

3 Russell FM, Greenwood B. Who should be prioritised for COVID-19 vaccination? Hum Vaccin Immunother 2021; 17: 1317–21.

4 Voysey M, Clemens SAC, Madhi SA, et al. Safety and efficacy of the ChAdOx1 nCoV-19 vaccine (AZD1222) against SARS-CoV-2: an interim analysis of four randomised controlled trials in Brazil, South Africa, and the UK. Lancet 2021; 397: 99–111.

5 Iacobucci G, Mahase E. Covid-19 vaccination: What’s the evidence for extending the dosing interval? BMJ 2021; 372: n18.

6 Department of Health, Northern Ireland. Introduction of shorter interval between vaccine doses | Department of Health. 2021; published online June 10. https://www.health-ni.gov.uk/news/introduction-shorter-interval-between-vaccine-doses (accessed Nov 25, 2021).

7 Davies NG, Kucharski AJ, Eggo RM, Gimma A, Edmunds WJ, Centre for the Mathematical Modelling of Infectious Diseases COVID-19 working group. Effects of non-pharmaceutical interventions on COVID-19 cases, deaths, and demand for hospital services in the UK: a modelling study. Lancet Public Health 2020; 5: e375–85.

8 Sandmann FG, Davies NG, Vassall A, Edmunds WJ, Jit M, Centre for the Mathematical Modelling of Infectious Diseases COVID-19 working group. The potential health and economic value of SARS-CoV-2 vaccination alongside physical distancing in the UK: a transmission model-based future scenario analysis and economic evaluation. Lancet Infect Dis 2021; 21: 962–74.

9 Davies NG, Abbott S, Barnard RC, et al. Estimated transmissibility and impact of SARS-CoV-2 lineage B.1.1.7 in England. Science 2021; 372. DOI:10.1126/science.abg3055.

10 Davies NG, Barnard RC, Jarvis CI, et al. Association of tiered restrictions and a second lockdown with COVID-19 deaths and hospital admissions in England: a modelling study. Lancet Infect Dis 2021; 21: 482–92.

11 Barnard RC, Davies NG, Jit M, John Edmunds W. LSHTM: Updated roadmap assessment – prior to delayed Step 4, 7 July 2021 - GOV.UK. 2021. https://www.gov.uk/government/publications/lshtm-updated-roadmap-assessment-prior-to-delayed-step-4-7-july-2021 (accessed Sept 16, 2021).

12 Liu Y, Morgenstern C, Kelly J, Lowe R, CMMID COVID-19 Working Group, Jit M. The impact of non-pharmaceutical interventions on SARS-CoV-2 transmission across 130 countries and territories. BMC Med 2021; 19: 40.

13 Voysey M, Costa Clemens SA, Madhi SA, et al. Single-dose administration and the influence of the timing of the booster dose on immunogenicity and efficacy of ChAdOx1 nCoV-19 (AZD1222) vaccine: a pooled analysis of four randomised trials. Lancet 2021; 397: 881–91.

14 Manisty C, Otter AD, Treibel TA, et al. Antibody response to first BNT162b2 dose in previously SARS-CoV-2-infected individuals. Lancet 2021; 397: 1057–8.

15 Google. COVID-19 Community Mobility Reports. 2021. https://www.google.com/covid19/mobility/ (accessed May 18, 2021).

16 Liu Y, Sandmann FG, Barnard RC, et al. Optimising health and economic impacts of COVID-19 vaccine prioritisation strategies in the WHO European Region. medRxiv 2021; published online July 14. DOI:10.1101/2021.07.09.21260272.

17 Lopez Bernal J, Andrews N, Gower C, et al. Effectiveness of Covid-19 Vaccines against the B.1.617.2 (Delta) Variant. N Engl J Med 2021; 385: 585–94.

18 Callaway E. Delta coronavirus variant: scientists brace for impact. Nature 2021; 595: 17–8.

19 GISAID. GISAID - hCov19 Variants. 2021. https://www.gisaid.org/hcov19-variants/ (accessed Sept 17, 2021).

20 UK Government. Coronavirus vaccine - weekly summary of Yellow Card reporting. 2021. https://www.gov.uk/government/publications/coronavirus-covid-19-vaccine-adverse-reactions/coronavirus-vaccine-summary-of-yellow-card-reporting (accessed Sept 16, 2021).

21 Hippisley-Cox J, Patone M, Mei XW, et al. Risk of thrombocytopenia and thromboembolism after covid-19 vaccination and SARS-CoV-2 positive testing: self-controlled case series study. BMJ 2021; 374: n1931.

22 Flaxman A, Marchevsky NG, Jenkin D, et al. Reactogenicity and immunogenicity after a late second dose or a third dose of ChAdOx1 nCoV-19 in the UK: a substudy of two randomised controlled trials (COV001 and COV002). Lancet 2021; 398: 981–90.

23 Moghadas SM, Vilches TN, Zhang K, et al. Evaluation of COVID-19 vaccination strategies with a delayed second dose. PLoS Biol 2021; 19: e3001211.

24 Parry HM, Bruton R, Stephens C, et al. Extended interval BNT162b2 vaccination enhances peak antibody generation in older people. medRxiv 2021; published online May 17. DOI:10.1101/2021.05.15.21257017.

25 Appelman B, van der Straten K, Lavell AHA, et al. Time since SARS-CoV-2 infection and humoral immune response following BNT162b2 mRNA vaccination. EBioMedicine 2021; 72: 103589.

26 Yassi A, Grant JM, Lockhart K, et al. Infection control, occupational and public health measures including mRNA-based vaccination against SARS-CoV-2 infections to protect healthcare workers from variants of concern: A 14-month observational study using surveillance data. PLoS ONE 2021; 16: e0254920.

27 Romero-Brufau S, Chopra A, Ryu AJ, et al. Public health impact of delaying second dose of BNT162b2 or mRNA-1273 covid-19 vaccine: simulation agent based modeling study. BMJ 2021; 373: n1087.

28 Hill EM, Keeling MJ. Comparison between one and two dose SARS-CoV-2 vaccine prioritization for a fixed number of vaccine doses. J R Soc Interface 2021; 18: 20210214.

29 Nam A, Ximenes R, Yeung MW, et al. Modelling the impact of extending dose intervals for COVID-19 vaccines in Canada. medRxiv 2021; published online April 10. DOI:10.1101/2021.04.07.21255094.

30 Matrajt L, Eaton J, Leung T, et al. Optimizing vaccine allocation for COVID-19 vaccines shows the potential role of single-dose vaccination. Nat Commun 2021; 12: 3449.

