## Supplemental Material for "Dosing interval strategies for two-dose COVID-19 vaccination in 13 low- and middle-income countries of Europe: health impact modelling and benefit-risk analysis"

### Supplemental Tables

#### Table S1. Model Parameters

| Disease process characteristics | | |
|---|---|---|
| **Parameter** | **Value** | **Source** |
| Age-specific susceptibility ($u_i$) | 0.38 - 0.88 | Davies et al.[1] |
| Age-specific clinical progression rates ($y_i$) | 0.21 - 0.70 | Davies et al[1] |
| Age-specific infection fatality rates | Raw input: 5.2e-6 - 0.13<br>By age group: 6.7e-6 – 8.1e-2 | Levin et al.[2] |
| Age- and country-specific within-population contact pattern ($C_{i,j,0}$) | Country-specific | Prem et al.[3] |
| Country-specific population age structures | Country-specific | United Nations[4] |
| Relationship between mobility and population contact pattern | Defined by linear and nonlinear functions for the *workplace* and *other* settings, respectively. | Davies et al. by fitting to UK data[5] |
| Latent period ($1/\sigma$) | ~gamma (μ = 2.5, k = 2.5) | Pearson et al.[6]<br>Davies et al.[7]<br>Davies et al.[5]<br>Bi et al.[8]<br>Liu et al.[9]<br>Linton et al.[10]<br>Nishiura et al.[11] |
| Duration of preclinical infectiousness ($1/\gamma_p$) | ~gamma (μ = 1.5, k = 4) | |
| Duration of clinical infectiousness ($1/\gamma_c$) | ~gamma (μ = 3.5, k = 4) | |
| Duration of subclinical infectiousness ($1/\gamma_s$) | ~gamma (μ = 5, k = 4) | Assumed, consistent with Davies et al.[7] |
| Relative infectiousness of subclinical infections compared to clinical infections ($f$) | 0.5 | Assumed, consistent with Davies et al.[7] |
| Duration of infection-induced immunity ($1/\omega_n$) | 3 years | Hall et al.[12] |

| Mortality, mobility, and non-pharmaceutical intervention characteristics | |
|---|---|
| **Parameter** | **Source** |
| Country-level daily COVID-19 Mortality (including 7-day rolling average) | Ritchie et al.[13] |
| Observed country-specific community mobility | Google[14] |
| COVID-19 Government Response Stringency Index and Government Response Tracker by country | Hale et al.[15] |

| Vaccine characteristics | | |
|---|---|---|
| **Parameter** | **Value** | **Source** |
| First dose protection duration ($1/\omega_v$) | Baseline = 360 days<br>Sensitivity analysis: 120 days | Voysey et al.[16] |
| Vaccine roll-out scenarios | 0.03 by mid-2021 and 0.2 by end of 2021, 0.5 by end of 2022 | Gavi, the vaccine alliance[17]<br>World Health Organisation[18,19] |
| Maximum willingness to receive vaccination | 0.7 for those between 20-59 and 0.9 for those above 60 | Wouter et al.[20]<br>Robinson et al.[21]<br>UK Government[22] |

### Table S2. Model Description

| | | |
|---|---|---|
| $S_i(t)$ | Susceptible individuals among age group $i$ at time $t$ | |
| | $S_i(t+1) = S_i(t) \cdot (1 - \lambda_i(t)) - v_{1i}(t+1) \cdot \frac{S_i(t)}{P_{1i}(t)} + R_i(t) \cdot \omega_n$ | (1) |
| $V_{1i}(t)$ | Individuals among age group $i$ who received their first doses at time $t$ | |
| | $V_{1i}(t+1) = V_{1i}(t) \cdot (1 - \lambda_i(t) \cdot (1 - ve_i) - \omega_v) + v_{1i}(t+1) \cdot \frac{S_i(t)}{P_{1i}(t)} - v_{2i}(t+1) \cdot \frac{V_{1i}(t)}{P_{2i}(t)}$ | (2) |
| $S_{wi}(t)$ | Individuals among age group $i$ who received their first doses but the protection has waned at time $t$ | |
| | $S_{wi}(t+1) = S_{wi}(t) \cdot (1 - \lambda_i(t)) + V_{1i}(t) \cdot \omega_v - v_{2i}(t+1) \cdot \frac{S_{wi}(t)}{P_{2i}(t)}$ | (3) |
| $V_{2i}(t)$ | Individuals among age group $i$ who received their second doses at time $t$ | |
| | $V_{2i}(t+1) = V_{2i}(t) \cdot (1 - \lambda_i(t) \cdot (1 - v2e_i)) + v_{2i}(t+1) \cdot (\frac{S_{wi}(t)}{P_{2i}(t)} + \frac{V_{1i}(t)}{P_{2i}(t)})$ | (4) |
| $E_i(t)$ | Exposed individuals who are not protected by vaccines (either unvaccinated or have their first doses already waned) among age group $i$ at time $t$ | |
| | $E_i(t+1) = E_i(t) \cdot (1 - \sigma) + S_i(t) \cdot \lambda_i(t) + S_{wi}(t) \cdot \lambda_i(t)$ | (5) |
| $E_{vi}(t)$ | Exposed individuals who are protected by one dose of the vaccine among age group $i$ at time $t$ | |
| | $E_{vi}(t+1) = E_{vi}(t) \cdot (1 - \sigma) + V_{1i}(t) \cdot \lambda_i(t) \cdot (1 - ve_i)$ | (6) |
| $E_{v2i}(t)$ | Exposed individuals who are protected by one dose of the vaccine among age group $i$ at time $t$ | |
| | $E_{v2i}(t+1) = E_{v2i}(t) \cdot (1 - \sigma) + V_{2i}(t) \cdot \lambda_i(t) \cdot (1 - v2e_i)$ | (7) |

| | | |
|---|---|---|
| $I_{pi}(t)$ | Pre-clinical infectious individuals among age group $i$ at time $t$ | |
| | $I_{pi}(t+1) = I_{pi}(t) \cdot (1-\gamma_p) + E_i(t) \cdot \sigma \cdot y_i + E_{vi}(t) \cdot \sigma \cdot y_i \cdot (1-ve_d) + E_{v2i}(t) \cdot \sigma \cdot y_i \cdot (1-v2e_d)$ | (8) |
| $I_{ci}(t)$ | Clinical infectious individuals among age group $i$ at time $t$ | |
| | $I_{ci}(t+1) = I_{ci}(t) \cdot (1-\gamma_c) + I_{pi}(t) \cdot \gamma_p$ | (9) |
| $I_{si}(t)$ | Subclinical infectious individuals among age group $i$ at time $t$ | |
| | $I_{si}(t+1) = I_{si}(t) \cdot (1-\gamma_s) + E_i(t) \cdot \sigma \cdot (1-y_i) + E_{vi}(t) \cdot \sigma \cdot (1 - y_i \cdot (1-ve_d)) + E_{v2i}(t) \cdot \sigma \cdot (1 - y_i \cdot (1-v2e_d))$ | (10) |
| $R_i(t)$ | Recovered individuals | |
| | $R_i(t+1) = R_i(t) \cdot (1-\omega_n) + I_{si}(t) \cdot \gamma_s + I_{ci}(t) \cdot \gamma_c - v_{1i}(t+1) \cdot \frac{R_i(t)}{P_{1i}}$ | (11) |
| $R_{vi}(t)$ | Individuals who have recovered from their previous infections and have received one dose | |
| | $R_{vi}(t+1) = R_{vi}(t) + v_{1i}(t+1) \cdot \frac{R_i(t)}{P_{1i}} - v_{2i}(t) \cdot \frac{R_{vi}(t)}{P_{2i}(t)}$ | (12) |
| $R_{v2i}(t)$ | Individuals who have recovered from their previous infections and have received two doses | |
| | $R_{v2i}(t+1) = R_{v2i}(t) + v_{2i}(t) \cdot \frac{R_{v2i}(t)}{P_{2i}(t)}$ | (13) |

In which:

| | |
|---|---|
| $\lambda_i(t)$ | Is the force of infection on population $i$ at time $t$: $$\lambda_i(t) = u_i \cdot \sum_{j=1}^{J} C_{i,j,t} \cdot \frac{(I_{pj}(t) + I_{cj}(t) + I_{sj}(t) \cdot f)}{N_j}$$ Where $j$ depicts age group, $J$ is 16, $f$ is the relative infectiousness of subclinical individuals compared to pre-clinical and clinical individuals (i.e. 50%), and $u_i$ is susceptibility. |
| $P_{1i}(t)$ | Is the population eligible for the first doses in the population $i$ at time $t$: $P_{1i}(t) = N_i - (V_{1i}(t) + S_{wi}(t) + R_{vi}(t) + E_{vi}(t)) - (R_{v2i}(t) + V_{2i}(t))$ |
| $P_{2i}(t)$ | Is the population eligible for the second doses in the population $i$ at time $t$: $P_{2i}(t) = V_{1i}(t) + S_{wi}(t) + R_{vi}(t) + E_{vi}(t)$ |
| $v_{1i}(t)$ | Is the number of doses to vaccinate the population with dose 1, this is pre-calculated based on vaccine dosing interval strategies. |
| $v_{2i}(t)$ | Is the number of doses to vaccinate the population with dose 2, this is pre-calculated based on vaccine dosing interval strategies. |

#### Table S3. The rationale behind vaccine effectiveness estimates

Vaccine effectiveness against pre-B.1.1.7 and B.1.1.7 relevant evidence (from roadmap report prior to step 4). This table is originally prepared for a report prepared by Barnard et al.[23]

| Description | Relevant evidence, assumed value shown in bold |
|---|---|
| Overall protection against infection for AstraZeneca dose 1 | Shrotri et al.[24] results, secondary analyses, paragraph 1, p.8 adjusted hazard ratio 0.33 (0.16, 0.68) at 28-34 days post vaccination. Pritchard et al.[25] supplementary Table 7, adjusted odds ratio >=21 days after first dose of AZ 0.36 (0.3, 0.45).<br><br>**0.67 (+28 days)** |
| Overall protection against disease for AstraZeneca dose 1 | Lopez Bernal et al.[26] Table 3, ChAdOx1 adjusted odds ratio d1:28-34 0.4 (0.27-0.59), adjusting 0.6 up to equivalent estimate for protection against infection (see cell above)<br><br>**0.67 (+28 days) as for infection** |
| Overall protection against hospitalisation for AstraZeneca dose 1 | Vasileiou et al.[27] Table 2, vaccine programme effect for ChAdOx1 21-27 days post first vaccine is 81% (72 to 87%), 28-34 days post first vaccine is 88% (75-94%), 35-41 days post first vaccine is 97% (63-100%). Smaller numbers. Table 3 splits analysis into age groups for ChAdOx1: 65-79 years 21-27 days post first dose 68% (31 to 85%), 80+ years 21-27 days post first dose 77% (63 to 86%) and 28-34 days post first dose 81% (60 to 91%). Small numbers for 65-79 years old and for 18-64 years old, so difficult to directly compare but overall the vaccine effect appears stronger in the younger (65-79 years) cohort than the older (80+) cohort, for the first three time points which enable comparison. Effect reversed for fourth time point. Ismail et al.[28] estimate vaccine effectiveness against hospitalisation of 73% (60-81%) for 80+ year olds and 84% (74-89%) for 70-79 year olds, 28 days following the first dose of AZ. When analysis is not split across vaccine products, the same study estimates efficacy against hospitalisation of 80% (74-85%) for 80+ year olds and 82% (75-87%) for 70-79 year olds.<br><br>**0.845 (+28 days)** |
| Overall protection against mortality for AstraZeneca dose 1 | Lopez Bernal et al.[29] estimated a hazard ratio of 0.45 (0.34 - 0.59) for cases vaccinated with one dose of AZ compared to unvaccinated cases, indicating an additional 55% (41-66%) protection against death given becoming a case for individuals vaccinated with one dose of AZ. Using the aforementioned estimate of a 55% increase and assuming this in addition to protection against |

| | |
|---|---|
| | disease of **0.67,** we get overall protection against mortality of 0.8515<br><br>**0.845 (+28 days)** |
| Overall protection against onward transmission for AstraZeneca dose 1 | Harris et al.[30] estimate an odds ratio of infection for contacts of index cases vaccinated with ChAdOx1 (AstraZeneca) matched to contacts of unvaccinated index cases of 0.62 (95% 0.48-0.79), where the vaccinated index cases received their vaccine dose at least 21 days before testing positive.<br><br>**0.38 (+28 days)** |
| Overall protection against infection for AstraZeneca dose 2 | Shrotri et al.[24] results, secondary analyses, paragraph 1, p.8 adjusted hazard ratio 0.32 (0.15, 0.66) at 35-48 days post vaccination<br><br>**0.68 (+14 days)** |
| Overall protection against disease for AstraZeneca dose 2 | Voysey et al.[31] A randomised controlled trial for ChAdOx1 nCoV-19 vaccine AZD1222, Table 3, average of efficacies more than 14 days after a second dose for LD/SD and SD/SD in 'COV002 (UK), age 18–55 years with >8 weeks' interval between vaccine doses*' row -> 0.778 = (0.9+0.656)/2<br><br>**0.78 (+14 days)** |
| Overall protection against hospitalisation for AstraZeneca dose 2 | Ismail et al.[28] estimate vaccine effectiveness against hospitalisation of 92% (87-95%) 14 days after a second dose across both AZ and Pfizer vaccines<br><br>**0.9 (+14 days)** |
| Overall protection against mortality for AstraZeneca dose 2 | **0.95 (+14 days)** |
| Overall protection against onward transmission for AstraZeneca dose 2 | **0.5 (+14 days)** |

### Supplemental Figures

#### Figure S1. Vaccine Roll-out Progress among LMICs of the WHO European Regions (n = 20)

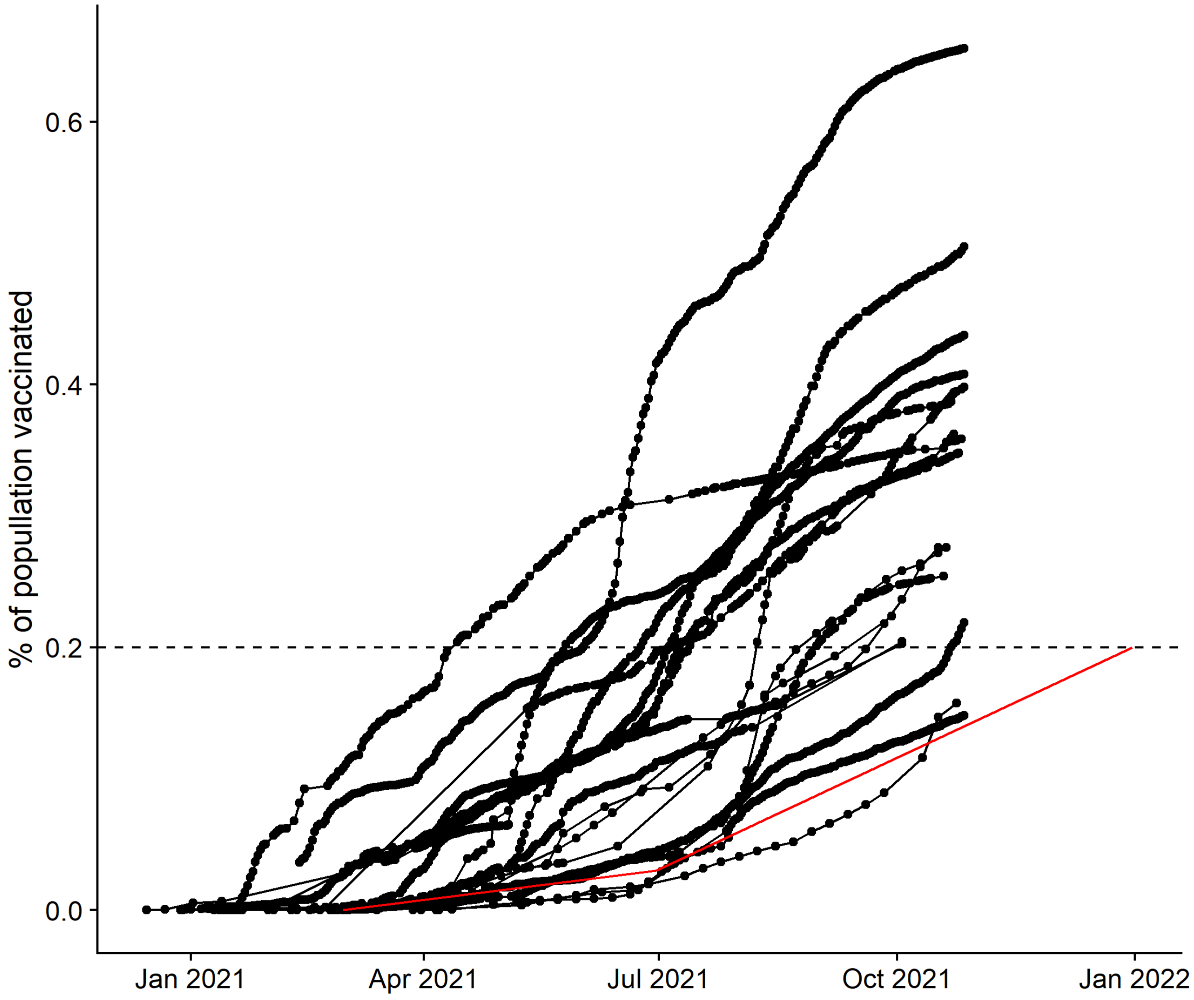

Caption: Each black dotted line represents a country; the red line represents the vaccine rollout strategy investigated in this study. Compared to the rest of the world, LMICs within the WHO European Region are relatively well off. We chose the slowest adaptor within the Region to resemble the slow vaccine row-out conditions that LMICs elsewhere in the world may face.

Figure S2. Outcomes using a dynamic relationship between dosing interval and the vaccine efficacy achievable after both doses.

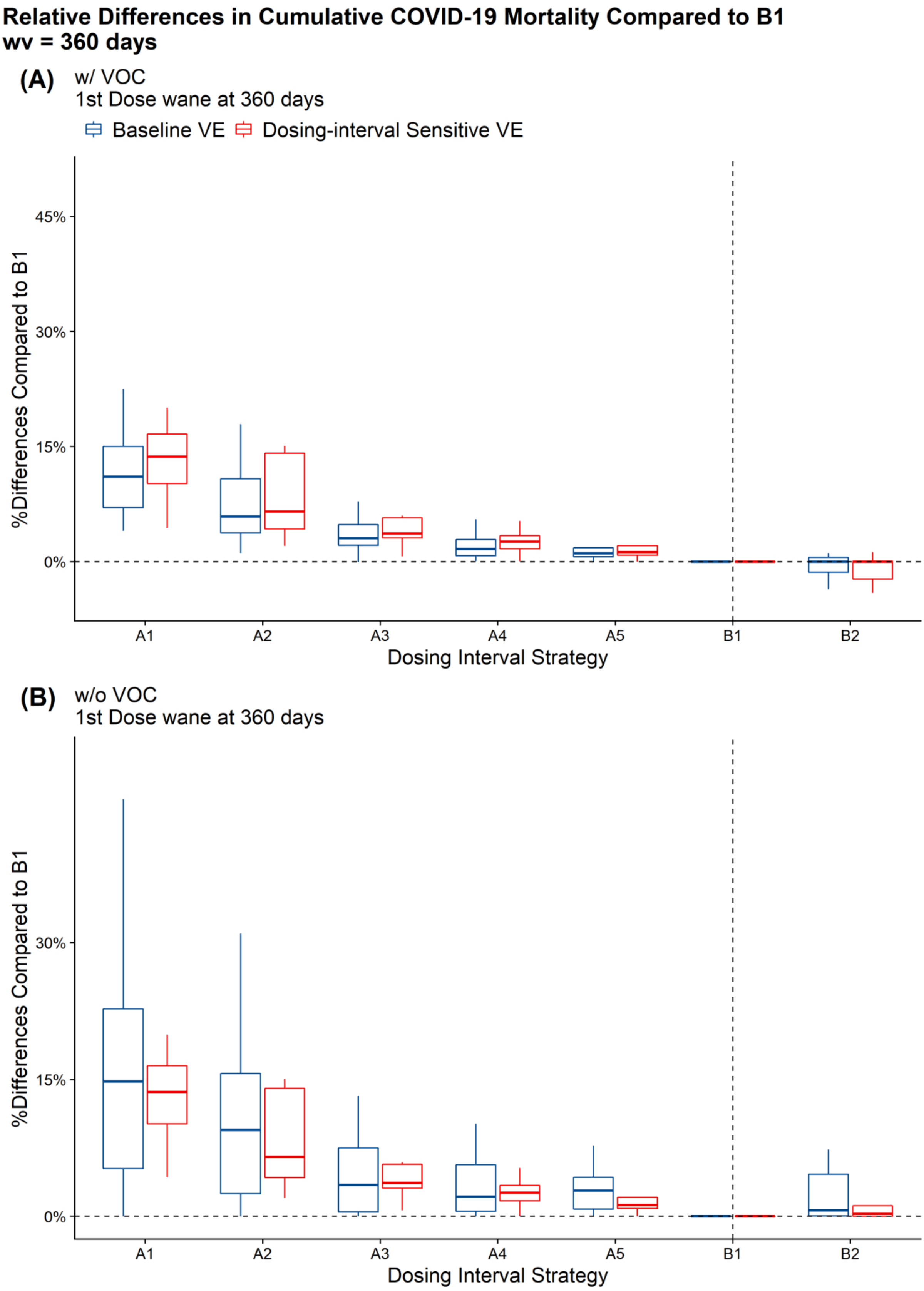

#### Figure S3. Sensitivity analyses around four dimensions of vaccine efficacy (infection- and disease-reducing vaccine efficacy after first and second doses).

The black dashed line shows 0% difference, the set of orange dashed lines show +/- 1% differences, and the set of red dashed lines show +/- 5% differences. All percentage differences were calculated using the COVID-19 mortality under strategy B1.

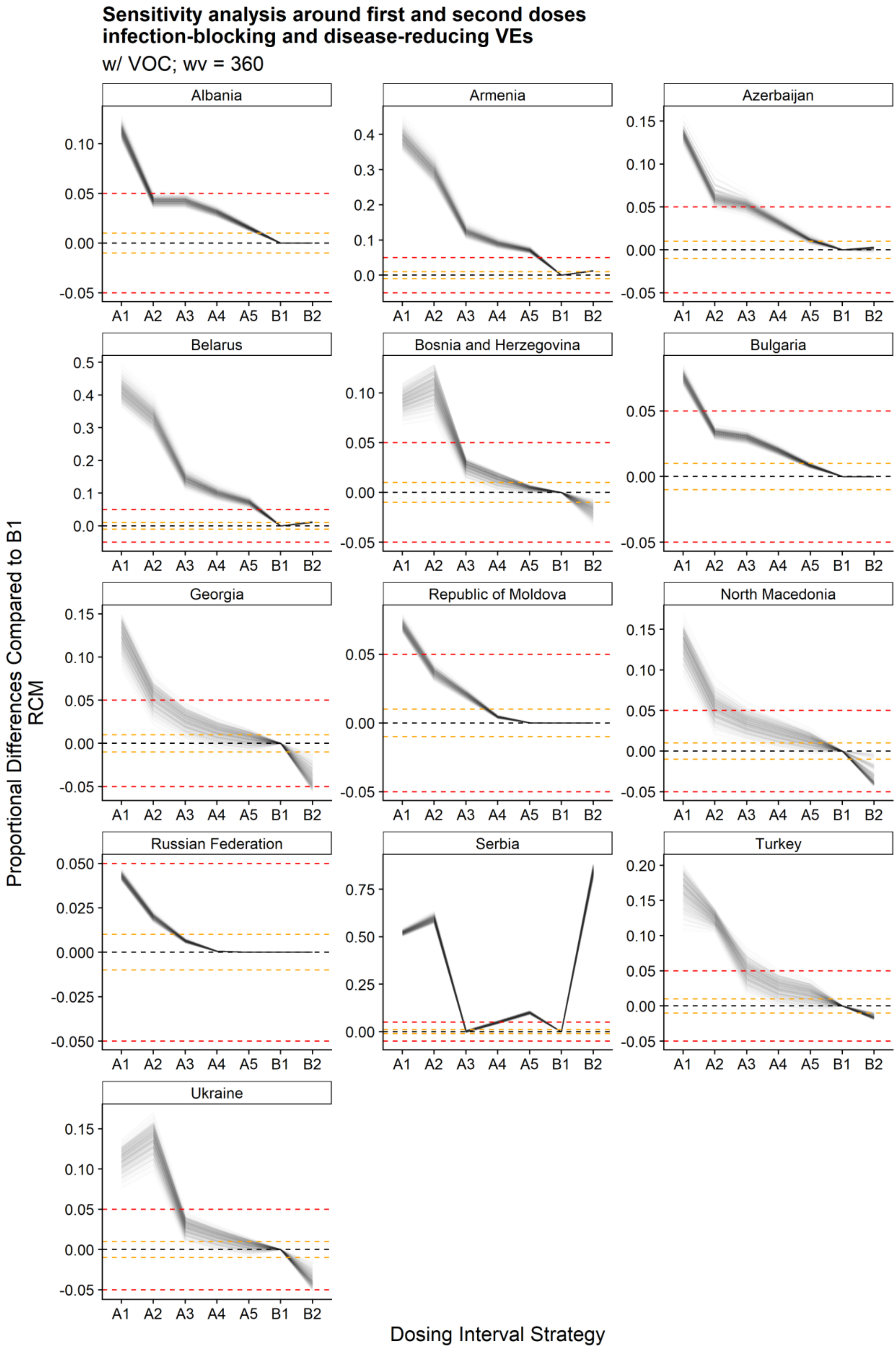
Sensitivity analysis around first and second doses infection-blocking and disease-reducing VEs
w/ VOC; wv = 360
Albania
Armenia
Azerbaijan
Belarus
Bosnia and Herzegovina
Bulgaria
Georgia
Republic of Moldova
North Macedonia
Russian Federation
Serbia
Turkey
Ukraine
A1 A2 A3 A4 A5 B1 B2
Proportional Differences Compared to B1 RCM
Dosing Interval Strategy

### Figure S4: Different outcomes by dosing strategy

Strategies A1-A5 and B1-B2 are arranged broadly based on mean effective dosing intervals. Results per strategy for each outcome are scaled to the highest value per country. Note that these values are relative measures to their respective public health measure – having higher relative measure in deaths than hospitalisations does not mean there are more deaths than hospitalisations, but only that the relatively changes in deaths compared to strategy A1 is greater.

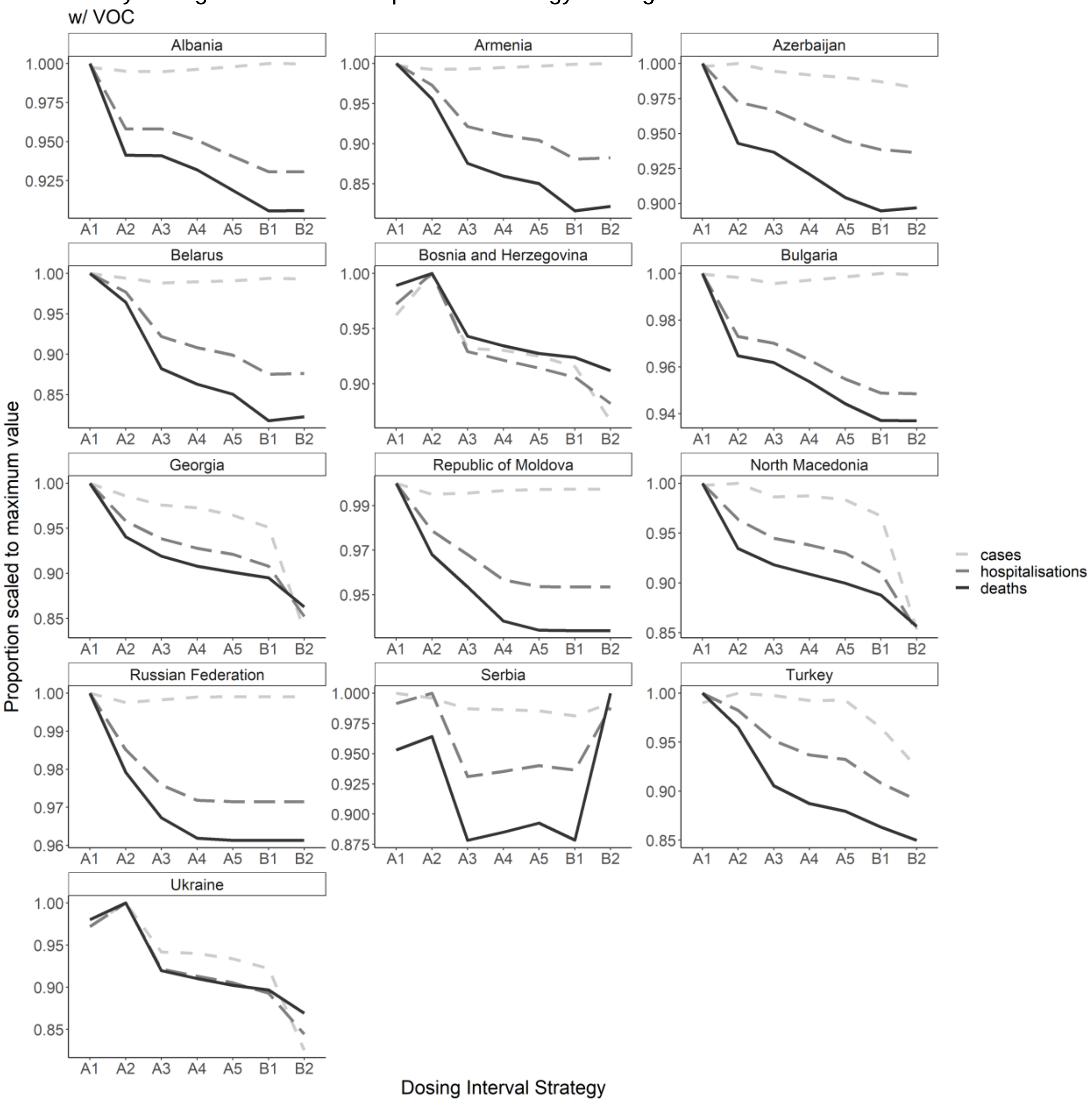

### Supplemental Methods

#### Calculating COVID-19 mortality and hospitalisation

This process has been previously described in Liu et al.

1. Infection to mortality delay function is assumed to be a gamma distribution with a mean of 26 (days) and a shape of 5. This probability density function is capped at 60 (days).[6]
$$P = Gamma(mean = 26; shape = 5)$$
2. The deaths that occurred on day $t$ due to infection on day $t - d$ can be expressed as (where $IFR_j$ depicts infection fatality ratio among age group $j$).
$$D(t)_{dj} = E(t-d)_j \cdot P(d) \cdot IFR_j$$
3. The total deaths occurred on day t can thus be expressed as:
$$D(t) = \sum_{j=1}^{J} \sum_{d=1}^{60} E(t-d)_i \cdot P(d) \cdot IFR_j$$

### Population contact patterns

We used observed community mobility to scale population contact patterns (`work` and `other` settings)[14] using  a set of parameters fitted as a part of another study(Davies et al. 2021) – significantly reduced community mobility is associated with relative reductions of contacts in the `work` and `other` settings. We used school-related non-pharmaceutical interventions recorded in the OxCGRT (Oxford Covid-19 Government Response Tracker) database to scale the contact patterns in the `school` setting[32] while keeping `home`-based contacts consistent with the pre-pandemic level.

While projecting into series not included in the fitting process, we assume the COVID-19 stringency index[32] (strictness of lockdown style policies that primarily restrict people's behaviour) would gradually return to a near-pre-pandemic level to capture potential long-term behaviour change. The community mobility and population contact patterns would return to near-pre-pandemic levels accordingly, using a Gaussian Generalized Additive Model (GAM) fitted to observations. Contacts in `school` settings follow school terms in the WHO European Region.

More technical details on the assumptions on the population contact patterns are described in Liu et al.[33]

### Setting up the sensitivity analyses around the vaccine efficacies

1. We created exhaustively the combinations of infection-reducing VE by first ($ve_i$) and second doses ($v2e_i$) and the disease-reducing VE for the first ($ve_d$) and second doses ($v2e_d$) by varying them between 0.25 and 0.95, with 0.1 increments; (4096 combinations)
2. We imposed rules that the vaccine efficacies of the first dose do not exceed the second dose (i.e., $ve_i \leq v2e_i$ and $ve_d \leq v2e_d$); (1296 combinations)
3. We imposed rules that the infection-blocking vaccine efficacies do not exceed the disease-reducing vaccine efficacies ($ve_i \leq ve_d$ and $v2e_i \leq v2e_d$); (540 combinations)

**Bibliography**

1 Davies NG, Klepac P, Liu Y, *et al.* Age-dependent effects in the transmission and control of COVID-19 epidemics. *Nat Med* 2020; **26**: 1205–11.

2 Levin AT, Hanage WP, Owusu-Boaitey N, Cochran KB, Walsh SP, Meyerowitz-Katz G. Assessing the age specificity of infection fatality rates for COVID-19: systematic review, meta-analysis, and public policy implications. *Eur J Epidemiol* 2020; **35**: 1123–38.

3 Prem K, Zandvoort K van, Klepac P, *et al.* Projecting contact matrices in 177 geographical regions: An update and comparison with empirical data for the COVID-19 era. *PLoS Comput Biol* 2021; **17**: e1009098.

4 United Nations. World Population Prospects 2019, United Nations Department of Economic and Social Affairs Population Division. 2019. https://population.un.org/wpp/ (accessed Nov 18, 2019).

5 Davies NG, Barnard RC, Jarvis CI, *et al.* Association of tiered restrictions and a second lockdown with COVID-19 deaths and hospital admissions in England: a modelling study. *Lancet Infect Dis* 2021; **21**: 482–92.

6 Pearson CAB, Bozzani F, Procter SR, *et al.* Health impact and cost-effectiveness of COVID-19 vaccination in Sindh Province, Pakistan. *medRxiv* 2021; published online Feb 25. DOI:10.1101/2021.02.24.21252338.

7 Davies NG, Kucharski AJ, Eggo RM, Gimma A, Edmunds WJ, Centre for the Mathematical Modelling of Infectious Diseases COVID-19 working group. Effects of non-pharmaceutical interventions on COVID-19 cases, deaths, and demand for hospital services in the UK: a modelling study. *Lancet Public Health* 2020; **5**: e375–85.

8 Bi Q, Wu Y, Mei S, *et al.* Epidemiology and transmission of COVID-19 in 391 cases and 1286 of their close contacts in Shenzhen, China: a retrospective cohort study. *Lancet Infect Dis* 2020; **20**: 911–9.

9 Liu Y, Centre for Mathematical Modelling of Infectious Diseases nCoV Working Group, Funk S, Flasche S. The contribution of pre-symptomatic infection to the transmission dynamics of COVID-2019. [version 1; peer review: 1 approved]. *Wellcome Open Res* 2020; **5**: 58.

10 Linton NM, Kobayashi T, Yang Y, *et al.* Incubation Period and Other Epidemiological Characteristics of 2019 Novel Coronavirus Infections with Right Truncation: A Statistical Analysis of Publicly Available Case Data. *J Clin Med* 2020; **9**. DOI:10.3390/jcm9020538.

11 Nishiura H, Linton NM, Akhmetzhanov AR. Serial interval of novel coronavirus (COVID-19) infections. *Int J Infect Dis* 2020; **93**: 284–6.

12 Hall VJ, Foulkes S, Charlett A, *et al.* SARS-CoV-2 infection rates of antibody-positive compared with antibody-negative health-care workers in England: a large, multicentre, prospective cohort study (SIREN). *Lancet* 2021; **397**: 1459–69.

13 Ritchie H, Mathieu E, Rodés-Guirao L, *et al.* Coronavirus Pandemic (COVID-19). 2020; published online March 5. https://ourworldindata.org/covid-deaths (accessed April 11, 2021).

14 Google. COVID-19 Community Mobility Reports. 2021. https://www.google.com/covid19/mobility/ (accessed May 18, 2021).

15 Hale T, Angrist N, Goldszmidt R, *et al.* A global panel database of pandemic policies (Oxford COVID-19 Government Response Tracker). *Nat Hum Behav* 2021; **5**: 529–38.

16 Voysey M, Costa Clemens SA, Madhi SA, *et al.* Single-dose administration and the influence of the timing of the booster dose on immunogenicity and efficacy of ChAdOx1 nCoV-19 (AZD1222) vaccine: a pooled analysis of four randomised trials. *Lancet* 2021; **397**: 881–91.

17 Gavi. The COVAX Facility: Interim Distribution Forecast – latest as of 3 February 2021. 2021. https://www.gavi.org/sites/default/files/covid/covax/COVAX-Interim-Distribution-Forecast.pdf (accessed March 26, 2021).

18 WHO. COVAX Announces additional deals to access promising COVID-19 vaccine candidates; plans global rollout starting Q1 2021. 2021. https://www.who.int/news/item/18-12-2020-covax-announces-additional-deals-to-access-promising-covid-19-vaccine-candidates-plans-global-rollout-starting-q1-2021 (accessed April 11, 2021).

19 WHO. COVAX reaches over 100 economies, 42 days after first international delivery. 2021. https://www.who.int/news/item/08-04-2021-covax-reaches-over-100-economies-42-days-after-first-international-delivery (accessed April 11, 2021).

20 Wouters OJ, Shadlen KC, Salcher-Konrad M, *et al.* Challenges in ensuring global access to COVID-19 vaccines: production, affordability, allocation, and deployment. *Lancet* 2021; **397**: 1023–34.

21 Robinson E, Jones A, Lesser I, Daly M. International estimates of intended uptake and refusal of COVID-19 vaccines: A rapid systematic review and meta-analysis of large nationally representative samples. *Vaccine* 2021; **39**: 2024–34.

22 Department of Health and Social Care (UK). UK COVID-19 vaccine uptake plan. 2021. https://www.gov.uk/government/publications/covid-19-vaccination-uptake-plan/uk-covid-19-vaccine-uptake-plan (accessed April 18, 2021).

23 Barnard RC, Davies NG, Jit M, John Edmunds W. LSHTM: Updated roadmap assessment – prior to delayed Step 4, 7 July 2021 - GOV.UK. 2021. https://www.gov.uk/government/publications/lshtm-updated-roadmap-assessment-prior-to-delayed-step-4-7-july-2021 (accessed Sept 16, 2021).

24 Shrotri M, Krutikov M, Palmer T, *et al.* Vaccine effectiveness of the first dose of ChAdOx1 nCoV-19 and BNT162b2 against SARS-CoV-2 infection in residents of Long-Term Care Facilities (VIVALDI study). *medRxiv* 2021; published online March 26. DOI:10.1101/2021.03.26.21254391.

25 Pritchard E, Matthews P, Stoesser N, *et al.* Impact of vaccination on SARS-CoV-2 cases in the community: a population-based study using the UK COVID-19 Infection Survey. *medRxiv* 2021; published online April 23. DOI:10.1101/2021.04.22.21255913.

26 Lopez Bernal J, Andrews N, Gower C, *et al.* Effectiveness of the Pfizer-BioNTech and Oxford-AstraZeneca vaccines on covid-19 related symptoms, hospital admissions, and mortality in older adults in England: test negative case-control study. *BMJ* 2021; **373**: n1088.

27 Vasileiou E, Simpson CR, Shi T, *et al.* Interim findings from first-dose mass COVID-19 vaccination roll-out and COVID-19 hospital admissions in Scotland: a national prospective

cohort study. *Lancet* 2021; **397**: 1646–57.

28 Ismail S, Vilaplana T, Elgohari S, *et al.* Effectiveness of BNT162b2 mRNA and ChAdOx1 adenovirus vectorCOVID-19 vaccines on risk of hospitalisation among older adults in England: an observational study using surveillance data. 2021. https://khub.net/documents/135939561/430986542/Effectiveness+of+BNT162b2+mRNA+and+ChAdOx1+adenovirus+vector+COVID-19+vaccines+on+risk+of+hospitalisation+among+older+adults+in+England.pdf/9e18c525-dde6-5ee4-1537-91427798686b (accessed July 15, 2021).

29 Bernal JL, Andrews N, Gower C, *et al.* Effectiveness of BNT162b2 mRNA vaccine and ChAdOx1 adenovirus vector vaccine on mortality following COVID-19. *medRxiv* 2021; published online May 18. DOI:10.1101/2021.05.14.21257218.

30 Harris R, Hall J, Zaidi A, Andrews N, Dunbar K, Dabrera G. Impact of vaccination on household transmission of SARS-COV-2 in England . 2021. https://khub.net/documents/135939561/390853656/Impact+of+vaccination+on+household+transmission+of+SARS-COV-2+in+England.pdf/35bf4bb1-6ade-d3eb-a39e-9c9b25a8122a?t=1619601878136 (accessed July 15, 2021).

31 Voysey M, Clemens SAC, Madhi SA, *et al.* Safety and efficacy of the ChAdOx1 nCoV-19 vaccine (AZD1222) against SARS-CoV-2: an interim analysis of four randomised controlled trials in Brazil, South Africa, and the UK. *Lancet* 2021; **397**: 99–111.

32 Blavatnik School of Government, University of Oxford. Oxford Covid-19 Government Response Tracker (OxCGRT). 2021. https://covidtracker.bsg.ox.ac.uk/ (accessed Sept 17, 2021).

33 Liu Y, Sandmann FG, Barnard RC, *et al.* Optimising health and economic impacts of COVID-19 vaccine prioritisation strategies in the WHO European Region. *medRxiv* 2021; published online July 14. DOI:10.1101/2021.07.09.21260272.
